# Central Related Blood Stream Infection in Patients undergoing Hemodialysis in a tertiary care centre

**DOI:** 10.64898/2026.01.27.26344916

**Authors:** E Mahesh, Saravu Narahari Sourabha, Mohammad Yousuff, R Rajashekar, KC Gurudev, MS Gireesh, Pooja Prabhu

## Abstract

**Background:** Catheter-related bloodstream infection (CRBSI) is a major cause of morbidity and mortality among patients undergoing hemodialysis (HD), particularly in low- and middle-income settings where non-tunneled hemodialysis catheters (NTHC) are widely used. Local epidemiological data are essential to guide preventive and therapeutic strategies.

**Objectives:** To determine the prevalence, microbiological profile, antimicrobial resistance patterns, and clinical outcomes of CRBSI in patients undergoing HD via internal jugular NTHC at a tertiary care center in South India.

**Methods:** This retrospective observational study included adults initiated on HD using internal jugular NTHC between January 2017 and December 2023. Patients with pre-existing infections or catheters inserted elsewhere were excluded. CRBSI was defined using KDOQI criteria. Demographic, clinical, laboratory, microbiological, and outcome data were analyzed. Logistic regression identified risk factors, and receiver operating characteristic (ROC) analysis evaluated predictors of adverse outcomes.

**Results:** Among 396 patients (mean age 56.3 ± 14 years; 70.4% male), 65 (16.4%) developed CRBSI, with an incidence of 4.7 per 1000 catheter days. Emergency HD initiation (OR 14.86, p < 0.001) and access failure (OR 2.71, p = 0.004) significantly increased CRBSI risk, while planned initiation for uremic symptoms was protective. Patients with CRBSI had lower serum albumin and higher leukocyte counts. Gram-negative organisms predominated (53.8%), with *Klebsiella pneumoniae* being the most common isolate. High resistance was observed to β-lactam/β-lactamase inhibitor combinations and carbapenems. Gram-negative CRBSI was associated with significantly higher odds of hospitalization, ICU admission, inotropic support, and mortality. ROC analysis showed good predictive ability for adverse outcomes (AUC 0.73–0.77).

**Conclusions:** CRBSI remains a significant complication of NTHC-based HD. Predominant Gram-negative infections with high antimicrobial resistance are associated with worse clinical outcomes, underscoring the need for early permanent access creation, strict catheter care, and robust antibiotic stewardship.

## Introduction

Chronic Kidney Disease (CKD) contributes to significant mortality and morbidity in India (1). The prevalence of CKD ranges from 12-21% across different regions in India (1). Hemodialysis (HD) is the most common mode of renal replacement therapy (RRT) amongst CKD population in India, followed by renal transplantation (2). Majority of the patients are initiated on HD via non tunnelled HD catheters (NTHC) (3). Delayed initiation of HD, delayed creation of permanent HD access like arterio venous fistulas (AVF) and low socio economic status are few of the factors which lead to emergency initiation of HD via NTHC (3). Infections are the second most common cause of mortality amongst patients with CKD, following cardiovascular disease (3,4). NTHC are prone to get infected and can result in catheter related blood stream infection (CRBSI) (3,5). Factors than can potentially contribute to infection risk are method of catheter insertion, size of catheter insertion and duration in situ, administration of antibiotics via NTHC, impaired immunity due to CKD and other co morbid conditions, malnourishment and poor hygiene (4–6). Due to the widespread use of NTHC for initiation and continuation of HD till maturation of AVF, there is a need for local epidemiological data to guide infection control practices and improve outcomes. Hence, we conducted a retrospective study to determine the prevalence and micro biological profile of CRBSI in patients undergoing hemodialysis in a tertiary care centre in South India.

## Materials and Methods

This single centre, hospital based, retrospective observational study was conducted at M S Ramaiah group of hospitals. The study included all patients who were initiated of HD at our centre from January 2017-December 2023 via internal jugular vein (IJV) NTHC. We excluded all patients who had signs of infection prior to insertion of NTHC, and patients who had NTHC, inserted at other centres.

The aims and objectives of the study were to study the prevalence of CRBSI, clinico microbiological profile and the antibiotic resistance patterns of organisms causing CRBSI.

### Sample Size

Sample size is calculated using the formula: n = (Z2 x P x (1 - P))/e2 Where:

- Z = value from standard normal distribution corresponding to desired confidence level (Z=1.96 for 95% CI)
- P is expected true proportion
- e is desired precision (half desired CI width).

By considering the prevalence of CRBSI as 39.25%, minimum sample size required was 367.

### Definition

We followed National Kidney Foundation Kidney Disease Outcome Quality Initiative guidelines for defining CRBSI (7). We defined our patients as having definite and possible CRBSI.

#### Definite

Same organism from a semiquantitative culture of the catheter tip (>15 CFU/catheter segment) and from a BC in a symptomatic patient with no other apparent source of infection.

#### Possible

Defervescence of symptoms after antibiotic treatment or after removal of catheter in the absence of laboratory confirmation of BSI in a symptomatic patient with no other apparent source of infection.

### Blood Culture

Paired blood cultures were obtained (10ml each from peripheral venous line and catheter hub), under sterile conditions, inoculated in culture media (BACT/) and immediately transported to microbiology lab. Serum samples for complete blood count (CBC), liver function test (LFT), calcium and phosphorous were sent simultaneously. The empirical antibiotic policy for management of CRBSI at our center is intravenous (IV) Vancomycin 500mg and IV Amikacin 500mg stat followed once in 48 hours. If patients did not show any clinical improvement with respect to symptoms after 72 hours, catheter was removed. Antibiotics were changed as per culture and sensitivity report.

Data were entered in Microsoft Excel and analysed using IBM SPSS Statistics for Windows. Continuous variables were expressed as mean ± standard deviation (SD) and categorical variables as frequency (%). The Shapiro–Wilk test was used to assess normality of data distribution. Comparisons between patients with and without CRBSI were performed using the independent samples *t*-test for normally distributed continuous variables and the Mann– Whitney U test for non-normal variables. Categorical variables were compared using the Chi-square test or Fisher’s exact test where appropriate. Univariate logistic regression was applied to examine the association of serum albumin, total leukocyte count, and diabetes mellitus with CRBSI, and odds ratios (OR) with 95 % confidence intervals (CI) were reported. Receiver operating characteristic (ROC) curve analysis was performed to evaluate the predictive ability of Gram-negative infections for adverse clinical outcomes, and the area under the curve (AUC) was calculated to determine discriminative performance. We used Haldane Anscombe correction to give finite adjusted odd’s ratio with 95% CI for clinical outcomes in gram negative vs non-gram-negative organisms. A *p* value < 0.05 was considered statistically significant.

## Results

A total of 396 patients underwent NTHC insertion during the study period. Out of 396 patients, 279 (70.45%) were male and 117 (29.55%) were female. Mean age of study population was 56.33 ± 13.97 years.

Diabetic nephropathy was the most common native kidney disease amongst the study population with 269 patients (67.93%), followed by chronic interstitial nephritis in 50 patients (12.63%). IgA nephropathy was the most common primary glomerular disease leading to ESRD in our study population, with 17 patients (4.29%). Baseline characteristics are mentioned in figure 1.

**Figure 1:**
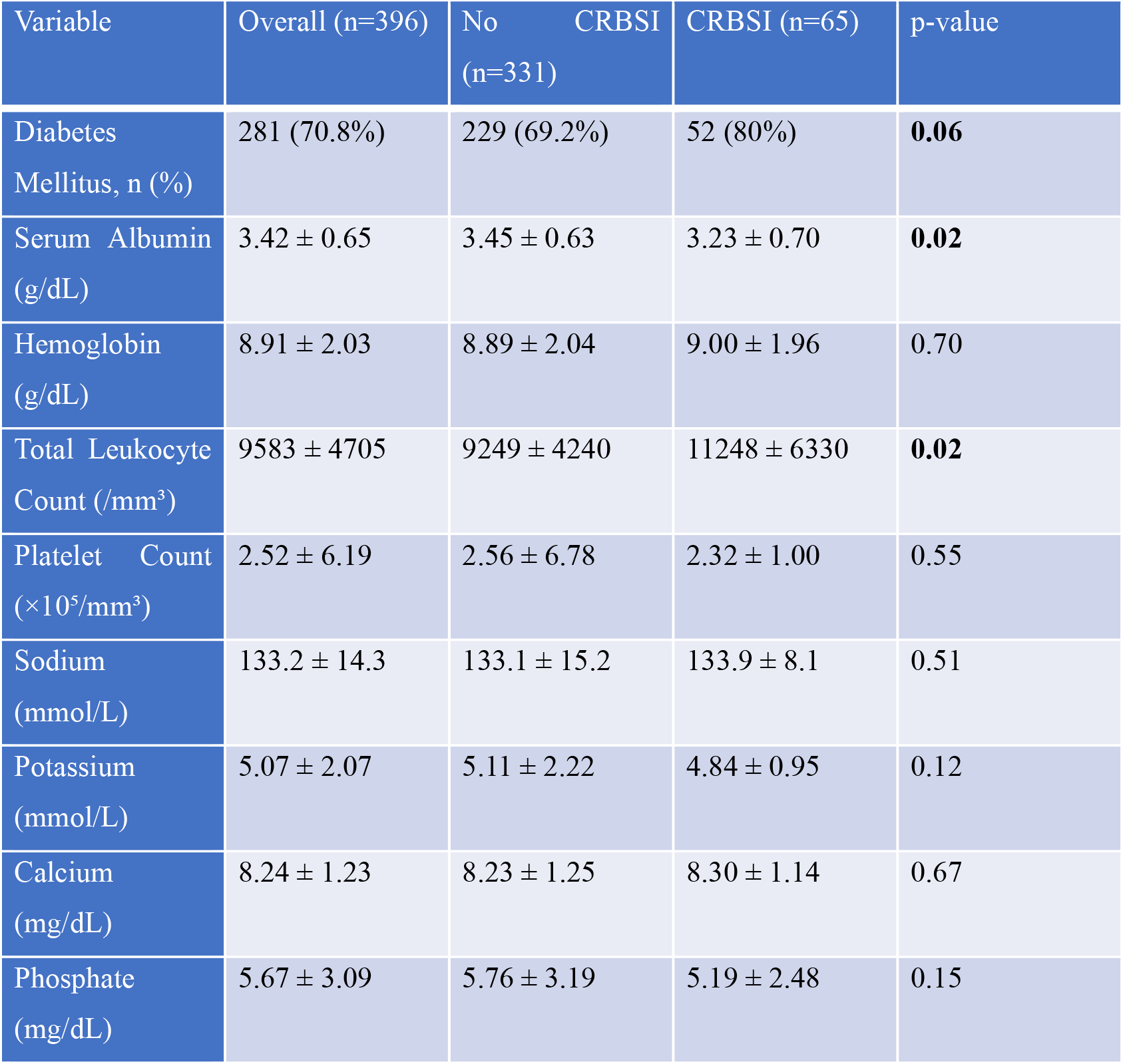
Baseline characteristics.

Diabetes mellitus was more common amongst patients with CRBSI as compared to those without CRBSI, however it was not statistically significant (p-0.06).

Patients with CRBSI had higher total counts (11248+6330) and lower serum albumin (3.23+0.70) as compared to non CRBSI group, reaching statistical significance (p-0.019, 0.02 respectively). On logistic regression modeling albumin as a continuous variable, each 0.5 g/dL decrease in serum albumin was associated with a 1.3-fold increase in odds of CRBSI (OR 1.32, 95% CI 0.94–1.85, p = 0.11). On logistic regression treating total leukocyte count as a continuous variable, each 1000 /mm^3^ increase was associated with an 8% higher odds of CRBSI (OR 1.08, 95% CI 1.03–1.14, p = 0.0037). On univariate logistic regression, the presence of diabetes mellitus increased the odds of CRBSI by 1.8-fold (OR 1.8, 95 % CI 0.94– 3.45, p = 0.076).

A total of 65 patients (16.4%) developed CRBSI. The incidence rate in our study was 4.7 infections per 1000 catheter days.

Patients who were initiated on HD on emergency basis (OR 14.86, 95% CI 4.63-47.68, p<0.001) and patients who had access failure (OR 2.71, 95% CI 1.31-5.60, p = 0.004) had significantly higher risk of developing CRBSI. In contrast, initiation of HD for uremic symptoms was less common in CRBSI group (OD 0.46, 95% CI 0.26-0.82, p = 0.015).

Klebsiella pneumoniae was the most common organism isolated from the culture, seen in 19 patients (29.2%), followed by Methicillin Resistant Staphylococcus Aureus (MRSA) and Serratia Marcesans being isolated in 6 patients each (9.23%). Enterobacter cloacae was isolated in 5 patients (7.7%). 22 patients had sterile cultures (33.8%) (Figure 3).

**Figure 2:**
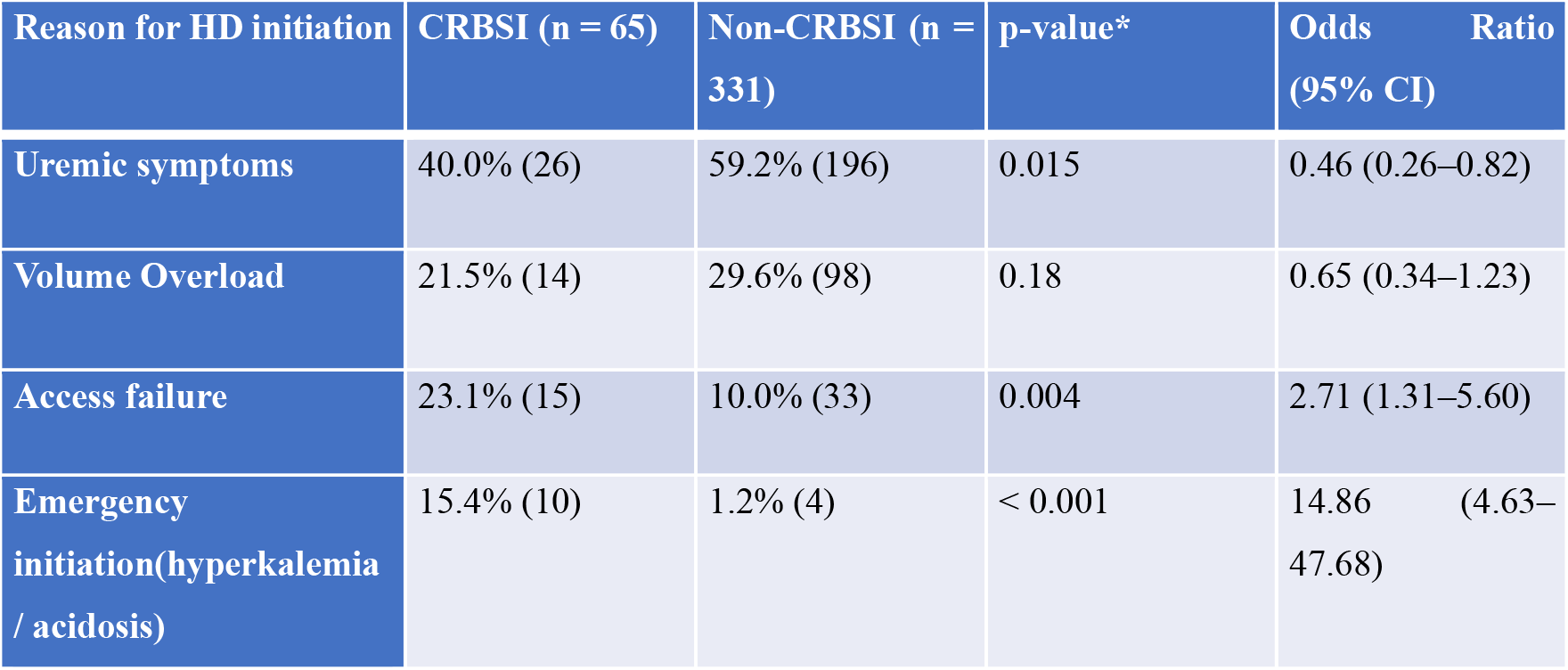
Reason for initiation of HD.

**Figure 3:**
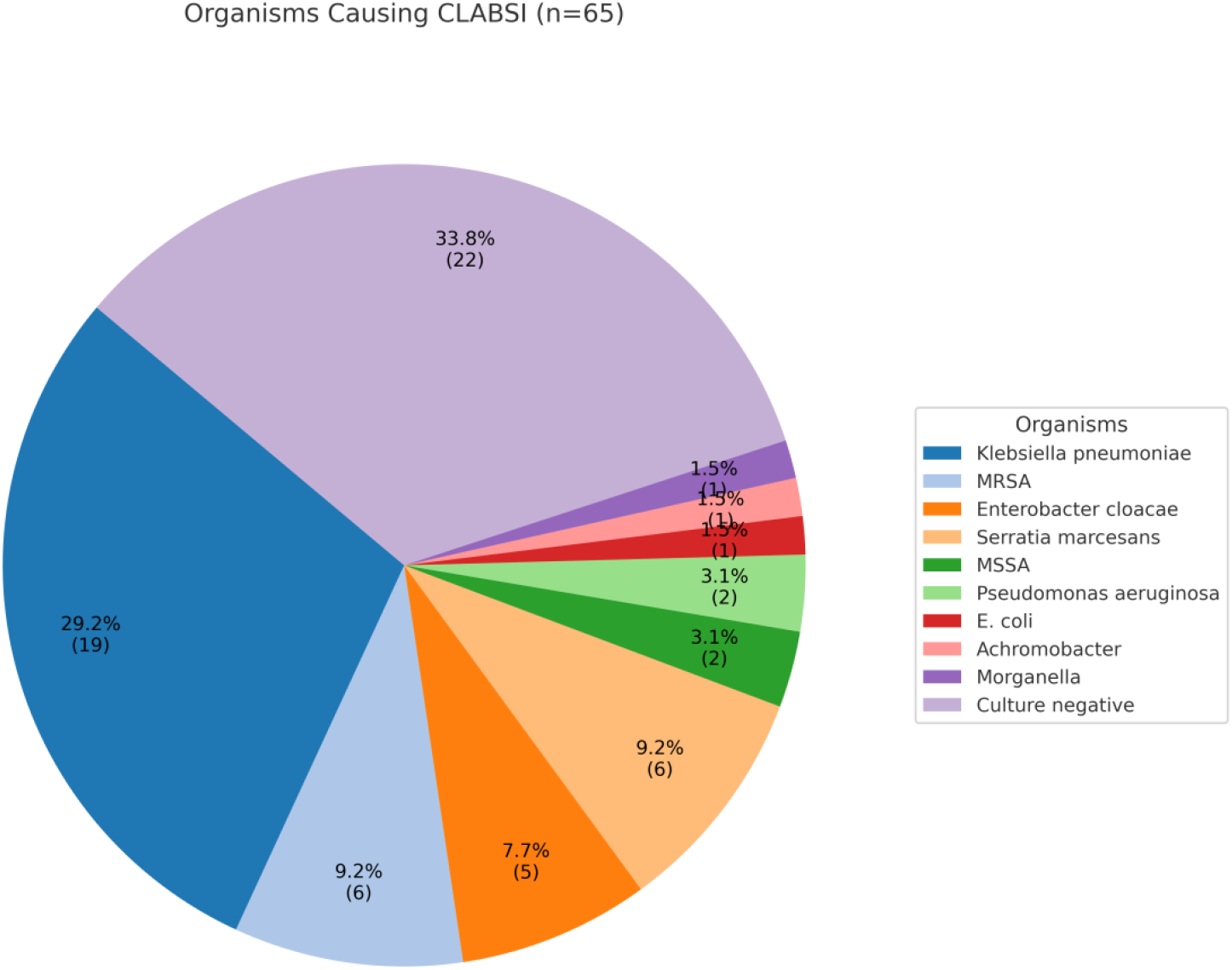
Organisms Causing CRBSI.

### Antibiotics resistance patterns

Gram negative organisms, Klebsiella pneumoniae being the most common, showed 84.2% resistance to both Piperacillin/Tazobactam and Cefaperazone+Sulbactum, 68% resistance to Meropenem, 63% resistance to Amikacin and 47% resistance to imipenem (Figure 4)

**Figure 4:**
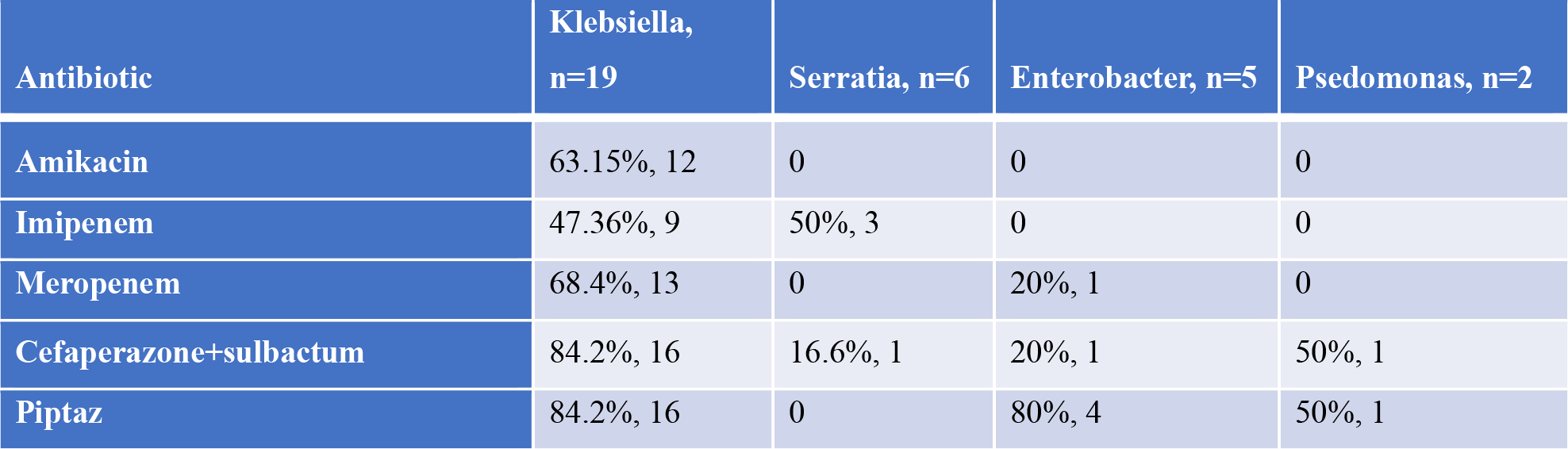
Antibiogram of Gram Negative CRBSI.

Serratia Marcesans showed 50% resistance to Imipenem and 16.6% resistance to Cefaperazone+Sulbactum, did not show any resistance to Amikacin or Piperacillin/Tazobactam. Enterobacter cloacae showed 80% resistance to Piperacillin/Tazobactam, 20% resistance to cefaperazone+sulbactum, however did not show any resistance to Amikacin. Pseudomonas aeruginosa showed 50% resistance to both piperacillin/tazobactam and cefaperazone+sulbactum, no resistance to amikacin (Figure 4)

MRSA isolates showed 66% resistance to levofloxacin, 50% to clindamycin and 16.6% to both vancomycin and teicoplanin. MSSA isolates were sensitive to all the antibiotics in the panel (Figure 5).

**Figure 5:**
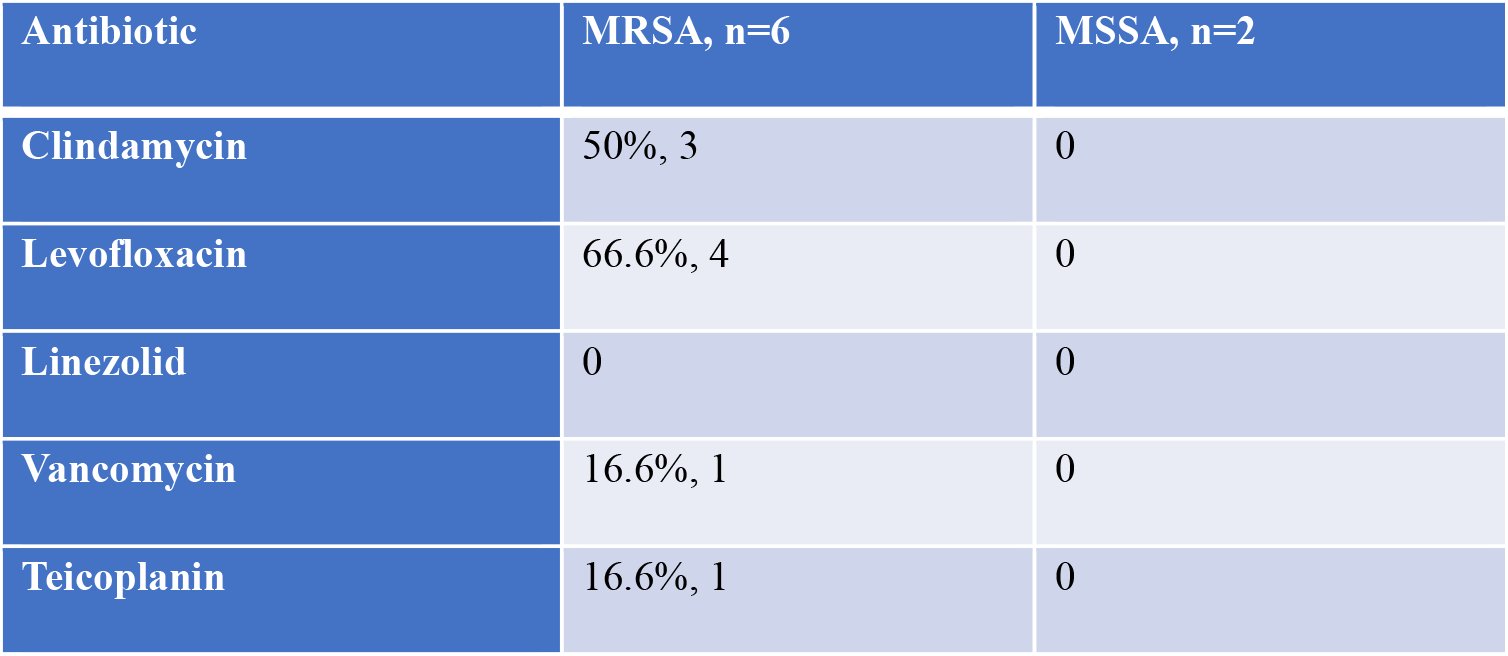
Antibiogram of Gram Positive CRBSI.

The median number of days at which all these patients developed CRBSI was 30.7+4.7 days.

### Clinical Outcomes

All patients received antibiotics for a mean duration of 8.05 days. Most common presenting complaints was fever with chills during hemodialysis (92%). 2 patients developed hypotension during HD, requiring immediate ICU care. Of the 65 NTHC, 27 patients (42%) had their catheters removed due to CRBSI. 38 (58%) patients did not have their NTHC removed. Majority of the patients (53.8%) had gram negative growth. Figure 6 depicts various clinical outcomes in all microbiological groups.

**Figure 6:**
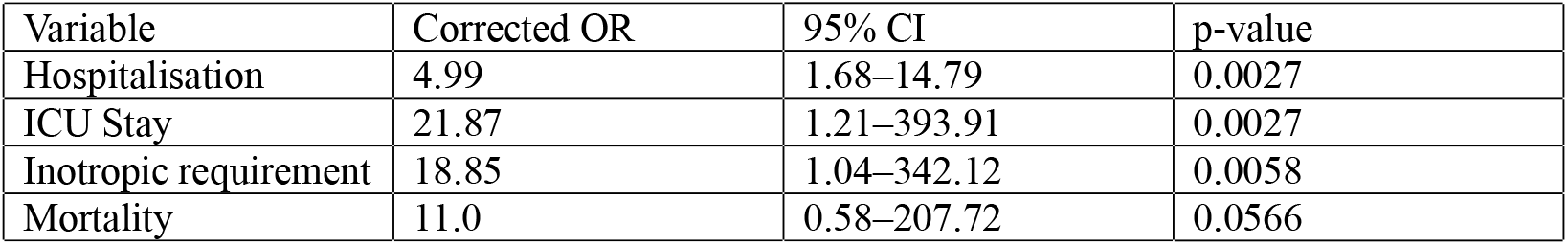
Clinical Outcomes in Microbiological Groups.

Gram negative infections were associated with 5 times higher odds of hospitalisation, 22 times higher chance of ICU stay and 19 times higher chance of requirement of inotropic support ( p = 0.0027, p = 0.0027, p = 0.005 respectively). Mortality showed 11 times higher OR with gram negative infection, but was not statistically significant (p = 0.056) (Figure 7)

**Figure 7:**
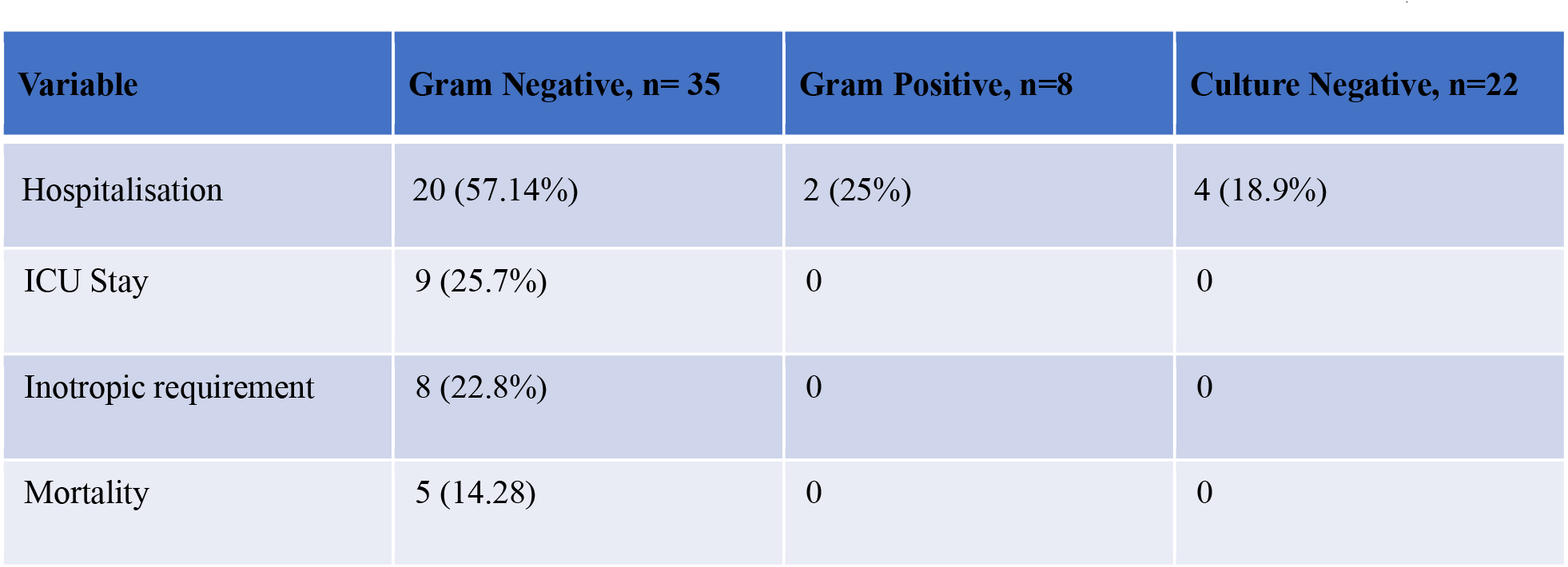
Corrected Odd’s Ratio for Gram Negative vs Non Gram Negative CRBSI.

Receiver operating characteristic (ROC) analysis demonstrated that Gram-negative infection predicted major adverse clinical outcomes with good discriminatory power (AUC range 0.73– 0.77). The highest AUCs were observed for inotropic support (0.77) and ICU stay (0.76), indicating that Gram-negative CRBSI is strongly associated with severe clinical course and critical care requirement. To improve estimate precision, a composite outcome (hospitalisation, ICU requirement, inotrope use, or mortality) was analysed. The composite severe outcome occurred in 53.6% of patients with Gram-negative CRBSI versus 11.1% of those with culture-negative infections (OR 9.23; 95% CI 2.3–37.3; *p* = 0.0011). (Figure 8, 9)

**Figure 8:**
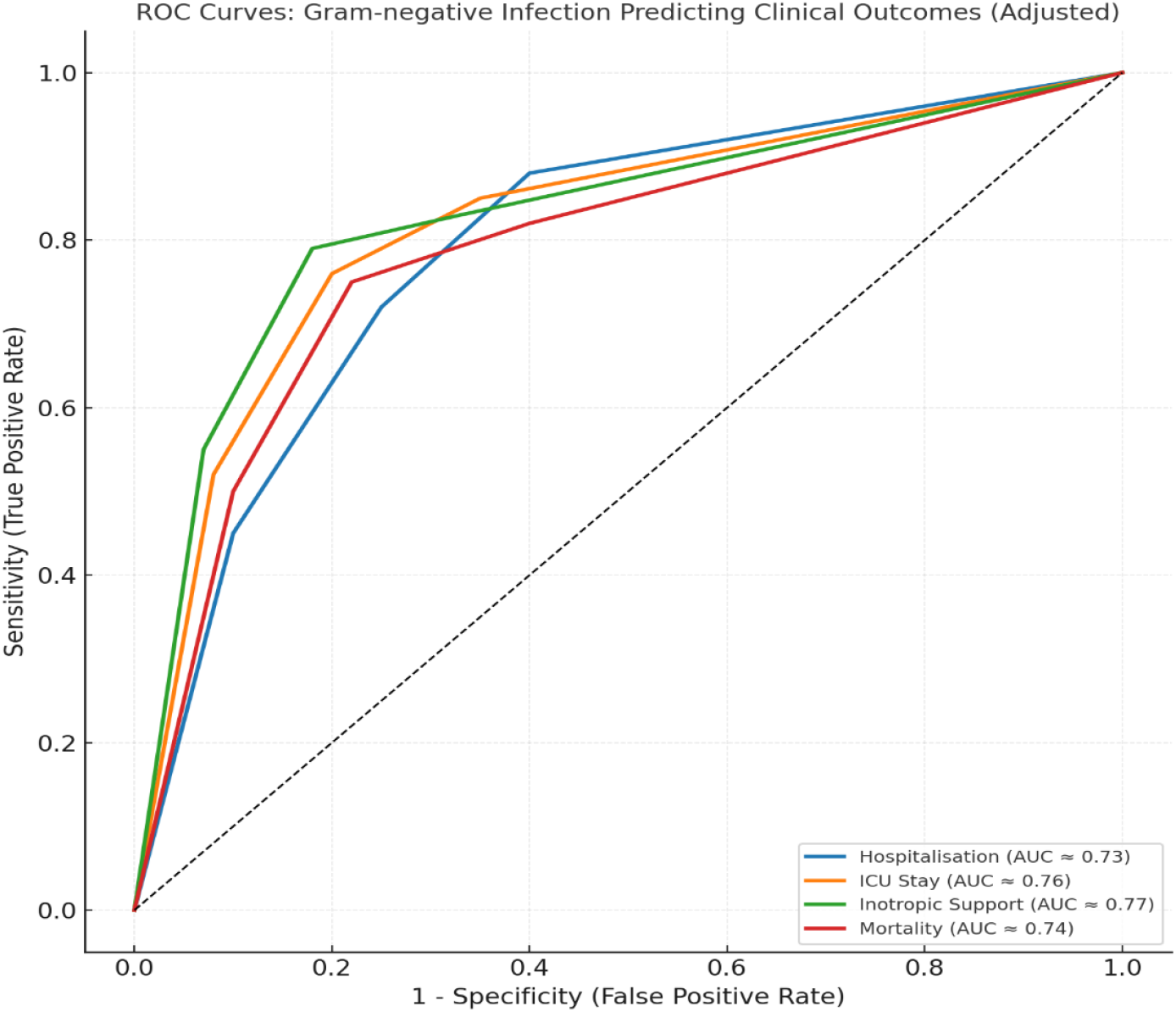
ROC curve depicting clinical outcomes with Gram Negative Organisms.

**Figure 9:**
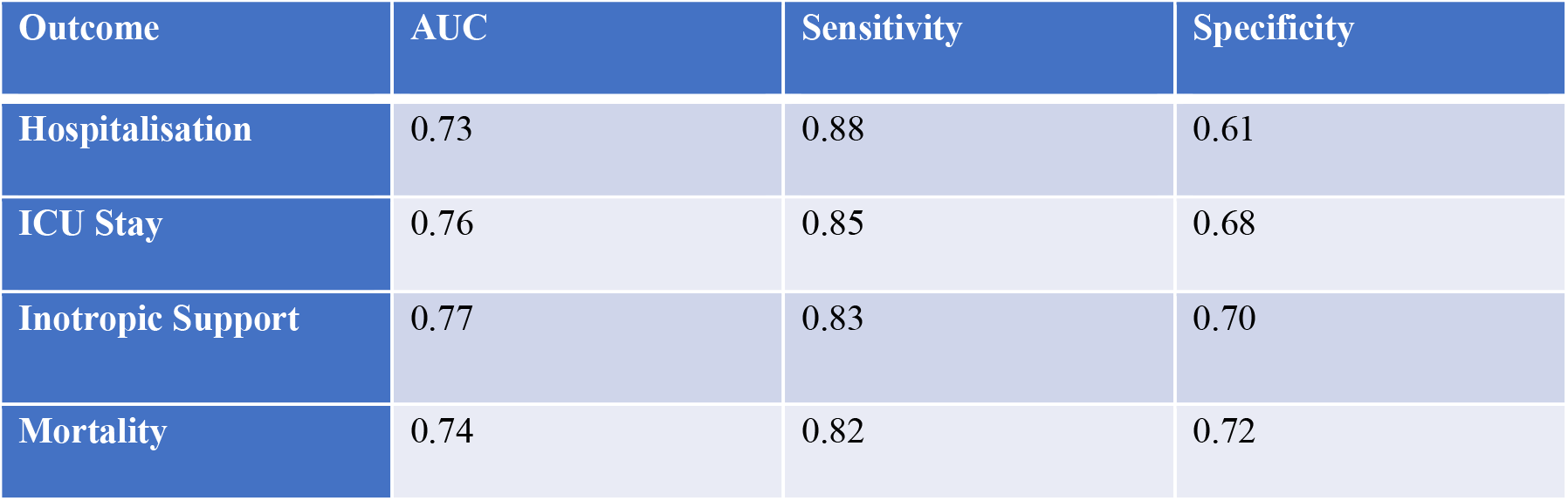
ROC Metrics.

## Discussion

Hemodialysis is the preferred modality of RRT in India due to various factors like financial constrain, lack of education (1,8). Majority of the patients would not have created an AVF or the fistula would not be matured yet to initiate the patient on HD via the same, hence they are initiated on HD via a NTHC. Due to delay in maturation or creation of AVF, mean duration of NTHC use is as high as 77 days (9). The mean duration of NTHC in situ in our study was 37.23 days, lower than various other studies (3,9).

70% of the study population consisted of males and diabetic nephropathy was the most common cause of ESRD. This is in concordance with study conducted by Lakshminarayana et al, R Hemachandar (8–10). The most common primary glomerular disease causing ESRD was IgA Nephropathy.

The prevalence of CRBSI in our study was 16.4%. This is much lower as compared to study conducted by Bhojaraja MV et al (23%), Schamroth Pravda et al (17.5%) and Opoku Asare et al (34%) (6,11,12). The use of HD catheter itself is a risk factor for developing infection and is almost three times higher as compared to AV graft and almost seven times higher as compared to an AVF (10). Hospitalisation for vascular access infection is also much higher for HD catheters as compared to AV grafts or AVF (10). The incidence of CRBSI in our study was 4.7 per 1000 catheter days for NTHC. KDOQI reports an incidence of 1.1 to 5.5 episodes of CRBSI per 1000 catheter days (7). The incidence of CRBSI in our study was much lower to studies conducted by Agarwal et al (7.4/1000), Bhojaraja MV et (13.38/1000) and P Pulampu et al (12.2/1000) (3,11,13). This lower incidence at our dialysis unit could due to adherence to aseptic precautions, good hand hygiene and lower mean duration of catheter placement (35 days). The median time to CRBSI in our study was 29.33 days, which is lower higher compared to studied conducted by Agarwal et al (24.5 days), Bhojaraja MV et al (17.2 days) (3,11).

Our study showed patients who were initiated on HD on emergency basis had higher risk of CRBSI (OR 14.86; 95% CI 4.63–47.68; *p* < 0.001), and patients who had access failure also had higher risk of CRBSI (OR 2.71; 95% CI 1.31–5.60; *p* = 0.004). The study by Lakshminarayana et al and R Hemachandar showed that 81% and 87% of the patients being initiated on HD on emergency basis via a NTHC, respectively (8,9). Similar to other studies, our study also showed emergency initiation of HD and patients with multiple access failures are at increased risk of having CRBSI (3,11). This may be attributed to the urgent nature of catheter insertion on emergency basis and limited adherence to aseptic precautions. In patients with multiple access failure, the goal is to preserve the access for as long as possible, hence the possibility of repeated catheter manipulation, prolonged catheter duration increases the likelihood of CRBSI. In contrast, patients who presented with uremic symptoms (OR 0.46; 95% CI 0.26-0.82 p = 0.015), had lesser incidence of CRBSI. This reflects a safer profile for planned, elective initiation of HD. These findings however, highlight the need for early AV access creation and patient education.

The majority of CRBSI in our study was caused by gram negative organisms (53.8%) with Klebsiella pneumoniae being the most common organism. 33.8% of our patients had probable CRBSI and 12.3% patients had gram positive organism. This correlates well with various other studies where gram negative organisms are the most common causative agent for CRBSI (3,4,12,14). Klebsiella was the most common gram negative isolate causing CRBSI in the study conducted by Schamroth Pravada et al (12). This is contrast to western data, where gram positive organisms were most commonly isolated (7,10,13). Gram positive bacteria usually reside on the colonizing areas around the HD catheter insertion site. Colonization leads to biofilm formation and acts as a nidus for bacteria. This difference is likely because of environmental and procedural factors like warmer climate, longer catheter dwell time and the higher incidence of nosocomial infections. Also, possibly high patient turnover in resource limited settings increases the risk of cross contamination with nosocomial gram negative organisms. Frequent use of broad spectrum antibiotics can possibly promote colonization and infection by gram negative organisms. Prolonged catheter duration and repeated catheter manipulation can increase exposure to hospital acquired flora, as seen in our study cohort.

Antibiogram data from our study showed that Klebsiella pneumoniae had 63% resistance to Amikacin, however the other gram negative organism did not show any resistance to amikacin. Klebsiella showed 84% resistance to both piperacillin/tazobactam and cefaperazone+sulbactum and 68% resistance to meropenem, whereas Enterobacter showed 80% resistance to piperacillin/tazobactam. Pseudomonas showed 50% resistance to both cefaperozone+sulbactum and pipercillin/tazobactam. Antibiogram data of gram positive organisms showed 16% resistance to both vancomycin and teicoplanin by MRSA. Local antibiogram is necessary to guide us for future therapy. Our study results were comparable in terms of gram negative organism drug susceptibility as compared to other Indian studies, however showed much higher vancomycin susceptibility (11,15). Our study showed high incidence of resistance to broad spectrum beta lactam/beta lactamase inhibitor combinations by Klebsiella species. Moreover, it showed high degree of resistance to carbapenems, an antibiotic usually reserved for patients with sepsis. The preserved activity of amikacin against other gram negative organism suggests that it still remains a viable empiric option. Our result highlights the need for periodic institutional antibiogram updates and antibiotic stewardship along with strict catheter care protocols.

In our study patients with CRBSI had significantly lower serum albumin levels and higher total counts, indication g the presence of chronic malnutrition due to poor nutritional status and chronic inflammatory state, both of which are associated with increased risk of blood stream infections, consistent with findings in other similar studies (3,11,16). Although diabetes mellitus was more prevalent in CRBSI group (80% vs 69%), the difference was not statistically significant (p=0.06). On univariate logistic regression, diabetes increased the odds of CRBSI by 1.8-fold (OR 1.80, 95% CI 0.94–3.45, *p* = 0.076). This correlates well with results seen in other studies (8–10,16). The pathogenesis might be attributable to microvascular lesions produced due to abnormal glucose metabolism resulting in tissue damage. Glucose rich environment causes proliferation of bacteria, leading to infection. Also, chronic hyperglycaemia can result in impaired immune response and dysregulation of macrophages. These findings highlight the multi factorial nature of risk of CRBSI.

Patients with gram negative infections more frequently required ICU admission, inotropic support and experienced mortality as compared to patients with non gram negative infections. In our study, gram negative infections had fivefold higher risk of hospitalisation (OR 4.99; 95% CI 1.68–14.79; p = 0.0027), 22-fold higher chance of ICU admission (OR 21.87; 95% CI 1.21– 393.91; p = 0.0027) and 19-fold greater requirement of inotropic support (OR 18.85; 95% CI 1.04–342.12; p = 0.0058). Although the odds of mortality were increased, it did not reach statistical significance (OR 11.0; 95% CI 0.58–207.72; p = 0.0566). Mortality occurred exclusively in the gram negative group. Composite severe outcome occurred in 53.6% of patients with Gram-negative CRBSI versus 11.1% of those with culture-negative infections (OR 9.23; 95% CI 2.3–37.3; p = 0.0011). ROC analysis (AUC 0.73-0.77) further confirmed that gram negative infections were moderate to strong predictors of adverse clinical outcomes, with sensitivity ranging from 0.82-0.88 and specificity ranging from 0.61-0.72. This is similar to the study by Agarwal et al and Bhojaraja et al where gram negative infections were associated with higher number of hospitalizations, requirement of ICU care and mortality (3,11).

Mortality observed in our study cohort was 7.7%, all caused by gram negative organisms (3 Klebsiella, 1 Pseudomonas and 1 Enterobacter). This is comparable to the studies by Agarwal et al and Bhojaraja et al, with mortality rates of 5.7% and 6.6% respectively (3,11).

## Data Availability

Data produced in present dtudy are available upon reasonable request to authors.

## References

1. Talukdar R, Ajayan R, Gupta S, Biswas S, Parveen M, Sadhukhan D, et al. Chronic Kidney Disease Prevalence in India: A Systematic Review and Meta-Analysis From Community-Based Representative Evidence Between 2011 to 2023. Nephrology. 2025 Jan;30(1):e14420.

2. Varughese S, Abraham G. Chronic Kidney Disease in India: A Clarion Call for Change. Clin J Am Soc Nephrol. 2018 May;13(5):802–4.

3. Agrawal V, Valson AT, Mohapatra A, David VG, Alexander S, Jacob S, et al. Fast and furious: a retrospective study of catheter-associated bloodstream infections with internal jugular nontunneled hemodialysis catheters at a tropical center. Clin Kidney J. 2019 Oct 1;12(5):737–44.

4. Pandit P, Sahni AK, Grover N, Dudhat V, Das NK, Biswas AK. Catheter-related blood stream infections: prevalence, risk factors and antimicrobial resistance pattern. Med J Armed Forces India. 2021 Jan;77(1):38–45.

5. Gahlot R, Nigam C, Kumar V, Yadav G, Anupurba S, Gahlot R, et al. Catheter-related bloodstream infections. Int J Crit Illn Inj Sci. 2014;4(2):162.

6. Opoku-Asare B, Boima V, Ganu VJ, Aboagye E, Asafu-Adjaye O, Asare AA, et al. Catheter-Related Bloodstream Infections among patients on maintenance haemodialysis: a cross-sectional study at a tertiary hospital in Ghana. BMC Infect Dis. 2023 Oct 7;23(1):664.

7. Lok CE, Huber TS, Lee T, Shenoy S, Yevzlin AS, Abreo K, et al. KDOQI Clinical Practice Guideline for Vascular Access: 2019 Update. Am J Kidney Dis. 2020 Apr;75(4):S1–164.

8. Lakshminarayana G, Sheetal L, Mathew A, Rajesh R, Kurian G, Unni V. Hemodialysis outcomes and practice patterns in end-stage renal disease: Experience from a Tertiary Care Hospital in Kerala. Indian J Nephrol. 2017;27(1):51.

9. Hemachandar R. Practice pattern of hemodialysis among end-stage renal disease patients in Rural South India: A single-center experience. Saudi J Kidney Dis Transplant. 2017;28(5):1150.

10. Hajji M, Neji M, Agrebi S, Nessira SB, Hamida FB, Barbouch S, et al. Incidence and challenges in management of hemodialysis catheter-related infections. Sci Rep. 2022 Nov 29;12(1):20536.

11. Bhojaraja MV, Prabhu RA, Nagaraju SP, Rao IR, Shenoy SV, Rangaswamy D, et al. Hemodialysis catheter-related bloodstream infections: a single-center experience. J Nephropharmacology. 2022 May 31;12(2):e10475.

12. Schamroth Pravda M, Maor Y, Brodsky K, Katkov A, Cernes R, Schamroth Pravda N, et al. Blood stream Infections in chronic hemodialysis patients - characteristics and outcomes. BMC Nephrol. 2024 Jan 3;25(1):3.

13. Puplampu P, Opoku-Asare B, Ganu VJ, Asafu-Adjaye O, Asare AA, Kyeremateng I, et al. Microbial organisms and antibiotic sensitivity patterns in patients with catheter-related bloodstream infections at a tertiary hospital. Clin Infect Pract. 2024 July;23:100365.

14. Emergence of antibiotic resistance in bloodstream infections associated with catheters in hemodialysis patients: a prospective observational study. J Ren Hepatic Disord [Internet]. 2025 [cited 2025 Oct 29]; Available from: https://jrenhep.com/index.php/jrenhep/article/view/211

15. Parameswaran R, Sherchan JB, Varma D M, Mukhopadhyay C, Vidyasagar S. Intravascular catheter-related infections in an Indian tertiary care hospital. J Infect Dev Ctries. 2010 Nov 11;5(06):452–8.

16. Guo H, Zhang L, He H, Wang L. Risk factors for catheter-associated bloodstream infection in hemodialysis patients: A meta-analysis. Shah A, editor. PLOS ONE. 2024 Mar 27;19(3):e0299715.

